# Sociodemographic Inequalities in Heat-Related Mortality across Summer Seasons from 2022 to 2024 in Peru

**DOI:** 10.1101/2025.01.31.25321450

**Authors:** Claudio Intimayta-Escalante

## Abstract

**Aim:** To assess sociodemographic inequalities in Heat-Related Mortality (HRM) across Peruvian departments during the summer seasons from 2022 to 2024.

**Methods:** An ecological study was conducted to analyze excess mortality during the summers of 2022–2023 and 2023–2024 in relation to maximum temperature records across 24 Peruvian departments. Sociodemographic factors, including sex, age group (*<*60 years or *≥*60 years), and educational level (elementary or lower vs. higher education), were considered. HRM was estimated for the overall population and stratified by sociodemographic characteristics using Poisson regression models. Inequality in HRM was quantified using the GINI index, where values near 1 indicate absolute inequality, while values closer to 0 suggest greater equality.

**Results:** A total of 70,832 deaths were analyzed, with 35,268 occurring in the summer of 2022–2023 and 35,564 in 2023–2024. The mean HRM was 29.24%, ranging from 2.26% to 166.67% across 10 departments. Higher HRM values were observed among females (HRM=51.90%), individuals *<*60 years (HRM=68.63%), and those with higher educational attainment (HRM=62.20%). The overall GINI index for HRM was 0.64, with greater inequality observed among females (GINI=0.38), individuals *<*60 years (GINI=0.51), and those with higher education levels (GINI=0.47).

**Conclusion:** Significant sociodemographic inequalities in HRM were observed across Peruvian departments during the summer seasons from 2022 to 2024. The highest inequalities were found among females, individuals *<*60 years, and those with higher educational attainment.

## INTRODUCTION

Climate change is assessed through multiple indicators, including temperature variations, precipitation patterns, sea level rise, and biodiversity loss.^1^ In last decades, global temperatures have risen, leading to more frequent and severe extreme heat events.^2^ Heat-related mortality (HRM) serves as a key indicator, as elevated temperatures can trigger an excess of deaths due to heat stroke, dehydration, and the exacerbation of chronic conditions, particularly among vulnerable populations such as children, the elderly, and individuals with pre-existing health conditions.^3,4^

Although Latin American countries have contributed minimally to anthropogenic climate change, they experience disproportionate impacts from extreme temperatures, particularly in low- and middle-income countries.^5^ However, the role of social, demographic, and economic factors in shaping HRM remains insufficiently explored.^6^ Peru, characterized by diverse geographic and climatic conditions, including variations in altitude, humidity, and population density, presents an ideal setting for examining these inequalities.^7^ Addressing these inequalities is crucial for understanding the broader impacts of extreme temperatures, which remain poorly comprehended.

Emerging evidence suggests that vulnerable populations are at greater risk of HRM, yet the extent of these inequalities across different regions is uncertain.^8^ Geographic, climatic, and social factors likely influence the unequal distribution of extreme temperature effects, underscoring the need for a comprehensive analysis. Therefore, this study aimed to assess sociodemographic inequalities in HRM across Peruvian departments during the summer seasons of 2022–2024.

## METHODS

### Study Design and Population

We conducted a longitudinal, retrospective, and ecological study estimating excess deaths during the summer seasons from December 2022 to March 2023 and from December 2023 to March 2024, across 24 Peruvian departments. The analysis included only deaths among the Peruvian population, excluding those due to violent causes (Appendix 1). In Peru, summer typically begins on December 21 and ends on March 20, bur exists regional variations in some departments, such as Lima—the capital which concentrated a third part of the population—experience more pronounced seasonal effects.^9^

### Sociodemographic Characteristics

Data on deceased individuals in Peru were obtained from the National Computerized System of Deaths or SINADEF in Spanish acronym (https://bit.ly/3PXyGBQ). This dataset provided information on key sociodemographic characteristics, including sex (male or female), age group (¡60 years or 60 years), and educational level (no education, primary, higher, or university). We calculated the total number of deaths during the summer periods and categorized them by these sociodemographic conditions.

### Heat-Related Mortality

Maximum temperature data for each department was obtained from meteorological stations managed by the National Service of Meteorology and Hydrology or SENAMHI in Spanish acronym (https://bit.ly/40RWnSh). We selected data from the stations closest to the capital of each department. So HRM was calculated using the following formula:

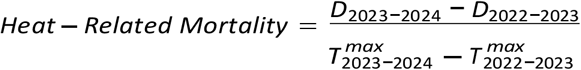

Where *D* represents the number of deaths across the 24 departments during the summer periods, and *T* denotes the maximum recorded temperature (°C) in each department for the corresponding periods.^10^

### Statistical Analysis

Statistical analyses were performed using R Studio version 4.2.2 (https://cran.r-project.org). In the descriptive analysis the frequencies and percentages were used for categorical variables, while mean with standard deviation (SD) were used for continuous variables. Group comparisons were conducted using the chi-square test for categorical data and the t-test for continuous data. To examine the association between positive excess deaths and rising maximum temperatures, we applied Poisson regression models, estimating the HRM as an extension of the Risk Ratio or RR with a 95% Confidence Interval (95%CI).^11^

### Inequality Analysis

Inequality was assessed using Lorenz curves to illustrate the uneven distribution of HRM across the accumulated population in different departments. The GINI index was then calculated, where values approaching 1 indicate absolute inequality, while values closer to 0 signify greater equality.^12^

### Ethical Aspects

This study was carried out using openly accessible data from the Peruvian population, which guaranteed confidentiality and anonymity. All analyses were carried out using aggregated departmental data while adhering to bioethical research principles.

## RESULTS

The analysis included 70,832 Peruvians who died in the summer periods of 2022-2023 (n=35,268) and 2023-2024 (n=35,564). Thus, no statistically significant differences were found according to sex (Table 1). However, there was a difference in the mean age of the deceased (69.63 and 70.46 years, for each summer season respectively). In addition, there were differences between educational levels for those deceased with no education or primary education (39.29% and 30.12%, respectively) and higher education or university (37.85% and 32.61%, respectively).

**Table 1.**
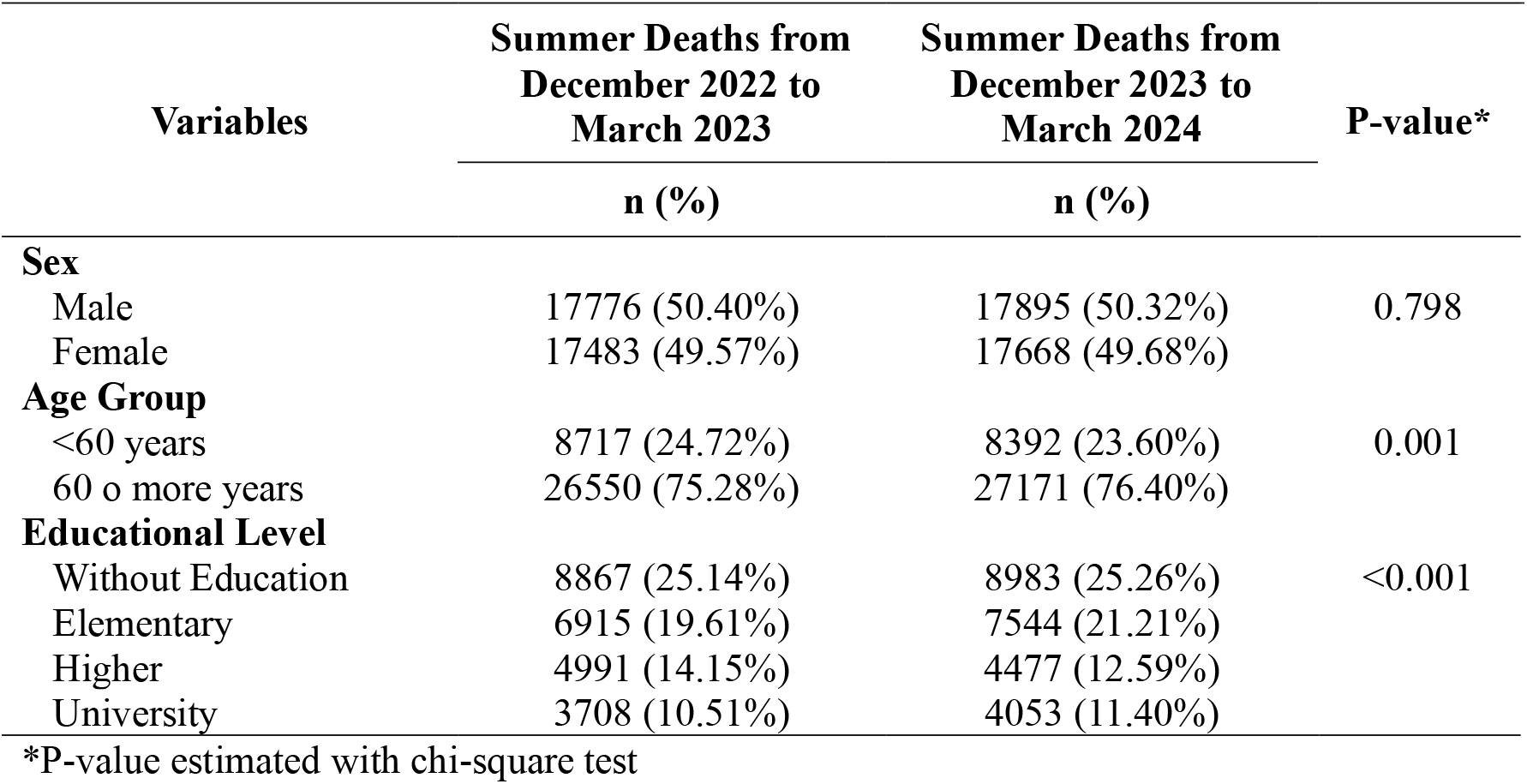
Differences in deaths of Peruvian population in summer seasons since 2022 to 2024.

The average of HRM was 29.24 (SD: 49.58), with values ranging from 2.26 to 166.67 in 10 Peruvian departments (Figure 1). In the evaluation of the average values according to sociodemographic characteristics, higher values were found in the female deceased population, those with¡60 years, and those with higher education or university (Table 2). These differences in HRM between the categories of characteristics evaluated were statistically significant.

**Table 2.** Average and Inequality of Heat-Related Mortality between Peruvian Departments in summer seasons since 2022 to 2024.

| Variables | Mean (95%CI) | P-value* | GINI (95%CI) |
| --- | --- | --- | --- |
| **Sex** |  |  |  |
| Male | 45.81 (28.11 – 63.52) | <0.001 | 0.38 (0.30 – 0.45) |
| Female | 51.90 (32.42 – 71.39) |  | 0.30 (0.14 – 0.40) |
| **Age Group** |  |  |  |
| <60 years | 68.63 (22.25 – 115.02) | <0.001 | 0.51 (0.28 – 0.62) |
| 60 o more years | 36.95 (3.84 – 70.07) |  | 0.43 (0.16 – 0.67) |
| **Educational Level** |  |  |  |
| Without Education or Elementary | 41.96 (15.61 – 68.31) | <0.001 | 0.43 (0.25 – 0.48) |
| Higher or University | 62.20 (29.95 – 94.45) |  | 0.47 (0.41 – 0.55) |

\*P-value estimated with the t-test

**Figure 1.**
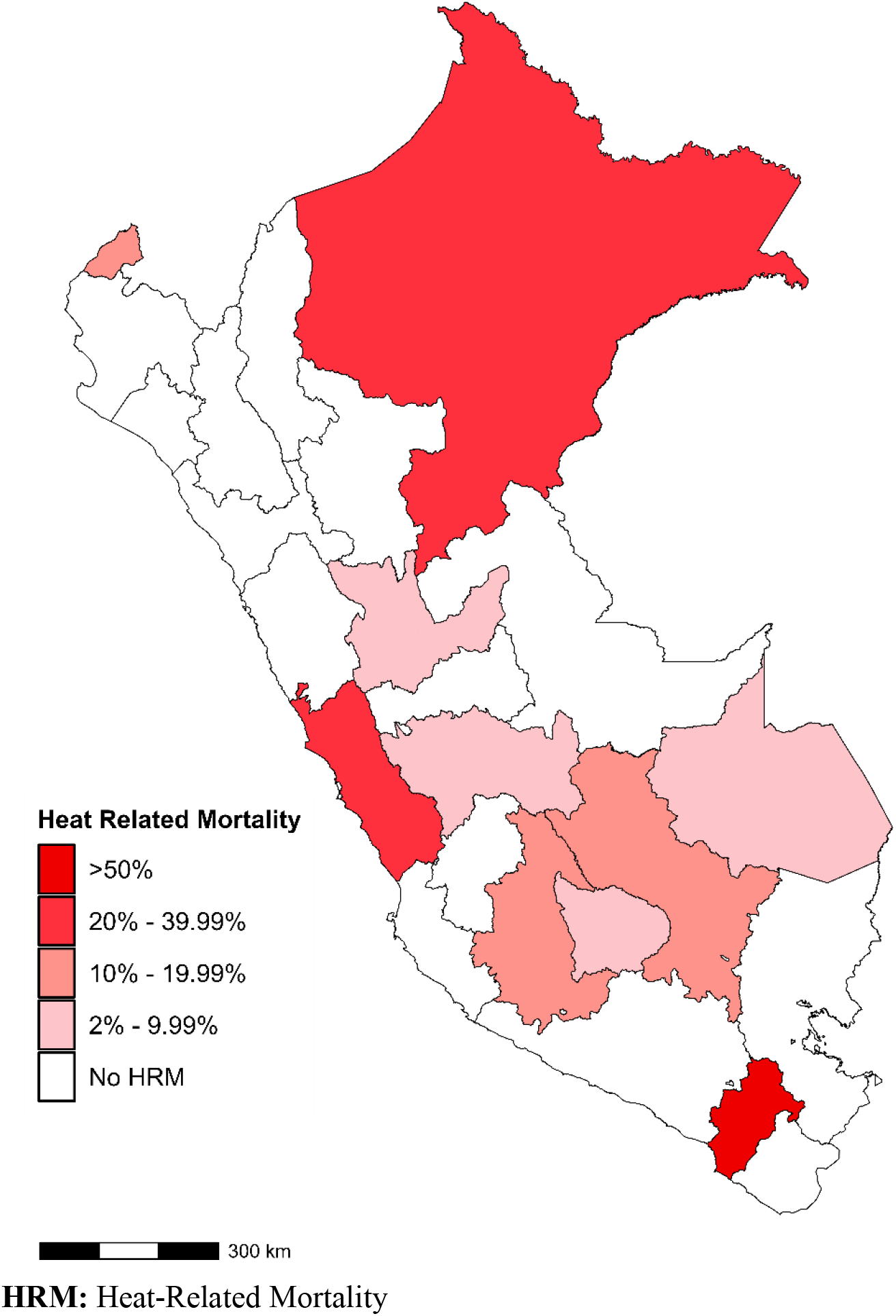
Geographical distribution of heat-related mortality between Peruvian departments in summer seasons since 2022 to 2024

In addition, the highest excess of HRM was estimated in five departments (Figure 2), with the highest in Moquegua (HRM=166.67%), followed by Loreto (HRM=34.07%), Lima (HRM=32.10%), Cusco (HRM=18.19%), and Tumbes (HRM=13.46%).

**Figure 2.**
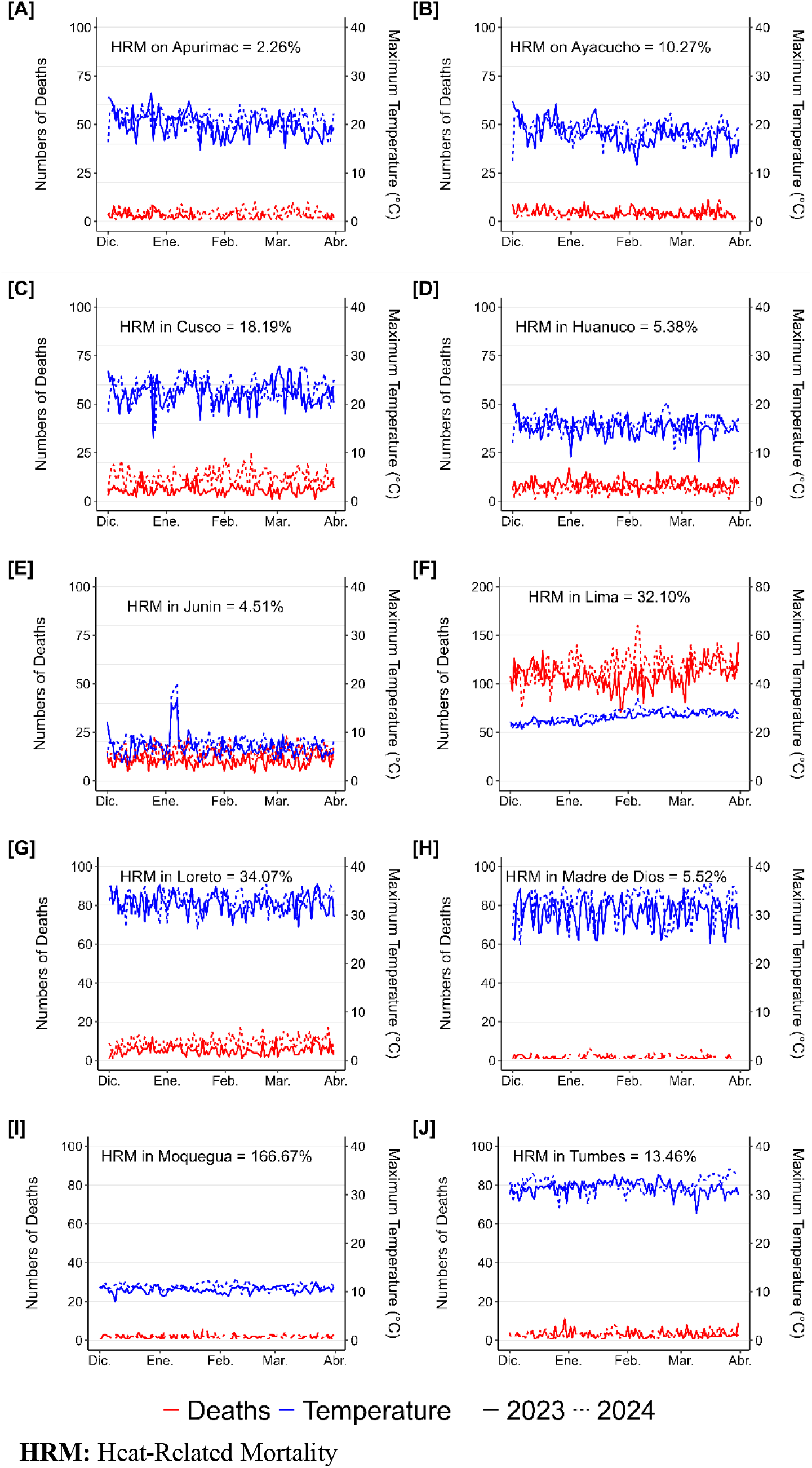
Excess deaths related to the increase in maximum temperature in Peruvian departments in summer seasons since 2022 to 2024

Inequality in HRM was estimated at a GINI of 0.64 (Figure 3). While the inequality levels were lower for various sociodemographic contexts, departments with a female deceased population (GINI=0.38), individuals with¡60 years old (GINI=0.51), and those with higher education or university (GINI=0.47) exhibited higher levels of inequality due to specific factors.

**Figure 3.**
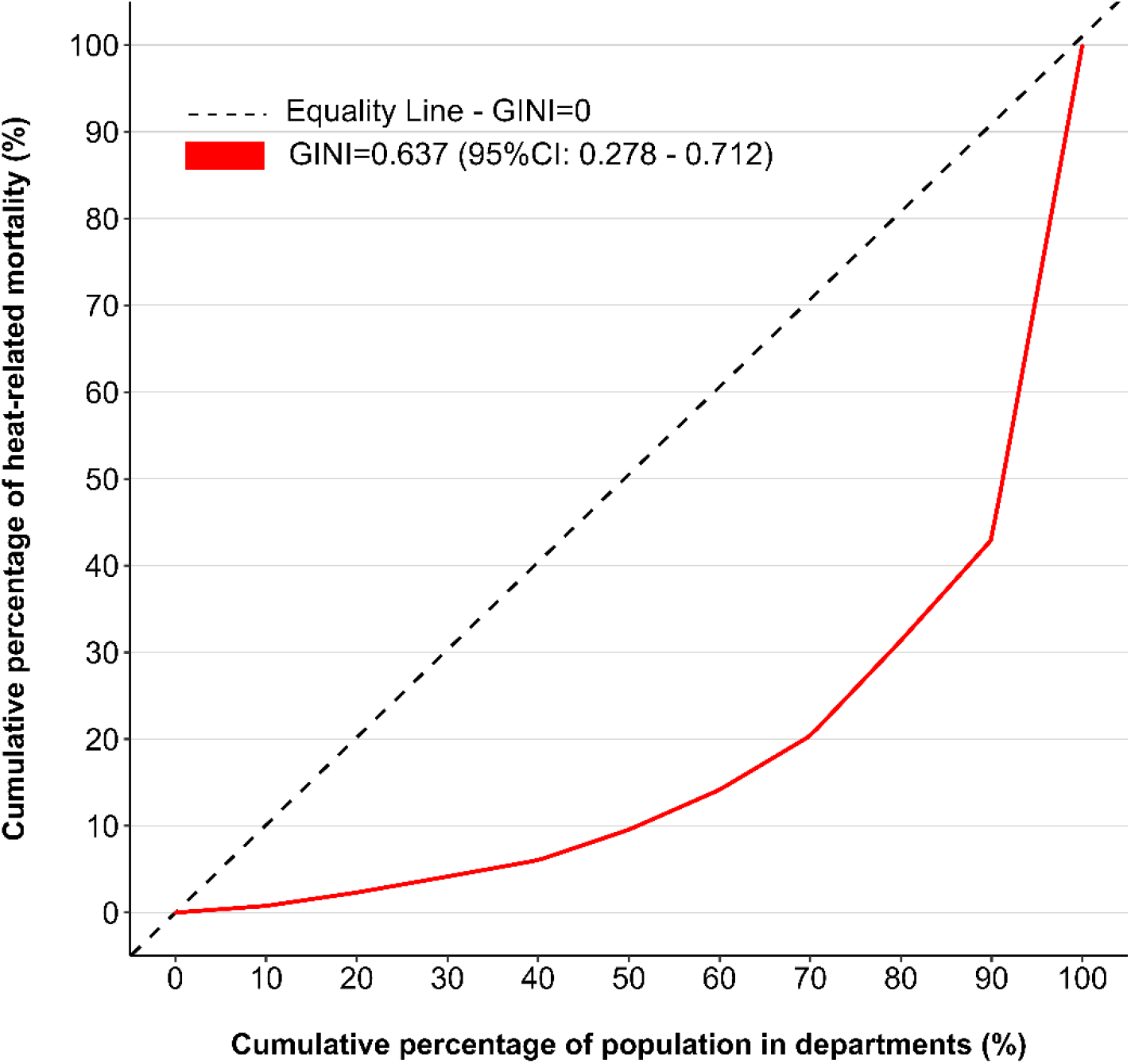
Inequality in heat-related mortality in Peruvian departments in summer seasons since 2022 to 2024

## DISCUSSION

This study assessed sociodemographic inequalities in HRM across Peruvian departments during the summer seasons from 2022 to 2024. Significant inequalities in HRM were observed for 10 departments, predominantly in the southern highland region, with a lesser impact in coastal areas. This pattern may be attributed to the moderating influence of the Peruvian sea on coastal temperatures, while high-altitude regions experience intensified solar radiation and lower humidity, exacerbating heat effects.^13,14^ HRM in Peru remains lower than in countries such as Brazil (166.12%), Paraguay (164.64%), and Bolivia (191.92%).^15^

Additionally, greater HRM inequalities were identified among females, individuals under 60 years old, and those with higher educational attainment. These inequalities may be driven by both biological and socioeconomic factors.^6,16^ In females, a higher proportion of body fat and lower muscle mass may alter thermoregulation and heat dissipation, increasing vulnerability to extreme temperatures.^17^ Individuals with higher education levels often reside in urban centers, where the urban heat island effect amplifies temperature extremes.^18^ Conversely, younger populations may experience heightened exposure due to precarious outdoor labor conditions.^19,20^

Unlike direct heat-related conditions such as heat stroke deaths, which are relatively rare in Peru, HRM captures excess of deaths linked to extreme temperatures variations without specifying cause of death.^21,22^ However, previous studies have documented the growing impact of climate change on Latin America, particularly in Brazil, Mexico, and Argentina, where vulnerable populations—especially the elderly and those with chronic illnesses—face heightened risks due to thermoregulatory challenges in extreme heat.^23,24^ Our findings align with this broader evidence, emphasizing the disproportionate burden of rising temperatures on socially and economically disadvantaged groups.

In contrast to prior research, this study not only estimates HRM, but also highlights the sociodemographic disparities in climate change-related mortality in Peru, which differs from other countries in the region.^25^ As adaptation and mitigation efforts against climate change advance in Latin America, future research should continue to evaluate the differential impacts of rising temperatures on various population groups.^26^ These insights can inform the development of targeted health interventions aimed at reducing climate-related health inequalities, particularly among vulnerable populations.

This study had strength in its rigorous evaluation of HRM and associated inequalities using specific statistical methods. However, several limitations should be acknowledged. First, the analysis was restricted to summer seasons after the COVID-19 pandemic periods in Peru, as temperature records for the entire year were unavailable across all departments. Second, the ecological design of the study limits extrapolation of finding at the individual level. Future research should incorporate longitudinal approaches to track exposure to extreme temperatures over time and explore additional sociodemographic and economic conditions of vulnerability related to health inequality.

In conclusion, this study provides evidence of significant sociodemographic inequalities in HRM across Peruvian departments during the summer seasons from 2022 to 2024. These inequalities were most pronounced among females, individuals under 60 years old, and those with higher educational attainment. The findings underscore the urgent need for targeted public health strategies to mitigate the rising impact of extreme temperatures on vulnerable populations.

## Data Availability

The data are available in open platforms in Peru, such as SINADEF (https://bit.ly/3PXyGBQ) or SENAMHI (https://bit.ly/40RWnSh).

https://bit.ly/40RWnSh

## Conflict of Interest

The author has no conflict of interest.

## Funding

This research was self-funded.

## APPENDICES

**APPENDIX 1**. Flow chart for the selection of the Peruvian deceased population

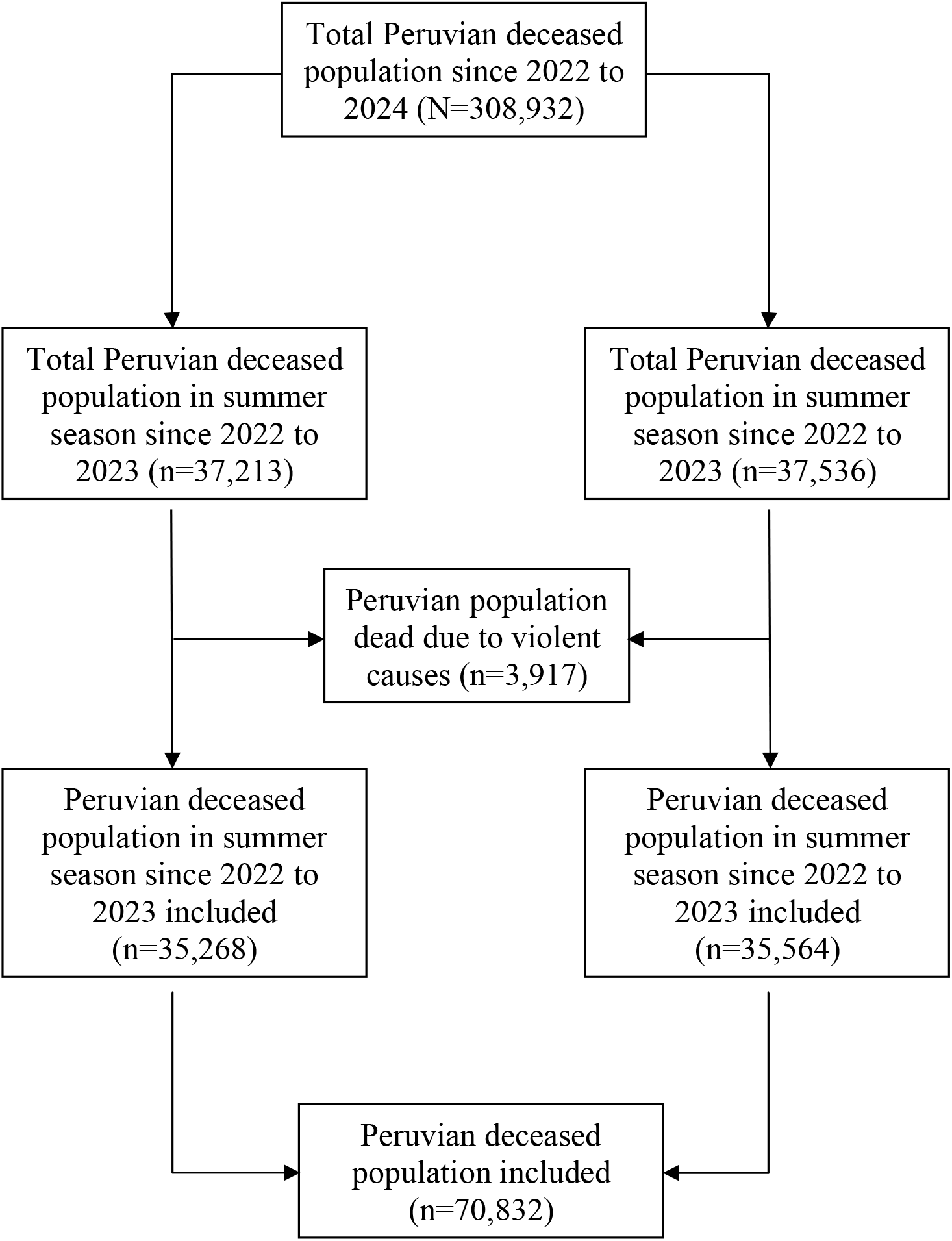

## Notes

### Competing Interest Statement

The authors have declared no competing interest.

### Funding Statement

This study was funded by the author

### Author Declarations

The study used ONLY openly available data

